# Antibody response to SARS-CoV-2 infection over six months among Nicaraguan outpatients

**DOI:** 10.1101/2021.04.28.21256122

**Authors:** Fredman González, Omar Zepeda, Christian Toval-Ruiz, Armando Matute, Hernan Vanegas, Nancy Munguia, Edwing Centeno, Yaoska Reyes, Lennart Svensson, Johan Nordgren, Aravinda M. de Silva, Sylvia Becker-Dreps, Lakshmanane Premkumar, Filemón Bucardo

## Abstract

New information is emerging about SARS-CoV-2 epidemiology and immunity, but little of this information comes from low- and middle-income countries or from patients receiving care in the outpatient setting. The current study investigated the SARS-CoV-2 infection status and antibody responses in 157 patients seeking care for a respiratory disease suggestive of COVID-19 in private healthcare clinics during the first wave (June–October 2020) of infections in Nicaragua. We examined nasal swabs for the presence of viral RNA via RT-PCR and longitudinally collected sera for the changes in SARS-CoV-2 Spike antibody levels over six months. Among patients with confirmed SARS-CoV-2 infections, we evaluated if clinical symptoms were associated with age, hematological parameters and co-morbidities. The combination of PCR and paired serology identified 60 (38%) of the 157 outpatients as acute COVID-19. While both PCR and serology identified the majority (n = 38, 64%) of the acute infections, a notable number of outpatients were identified by RT-qPCR (n = 13, 22%) or by serology (n = 9, 14%) only. During the longitudinal study, we identified 6 new infections by serology among the 97 non-COVID-19 subjects. In conclusion, this study report that more than one third of the outpatients seeking care for acute respiratory disease during the first epidemic wave of SARS-CoV-2 in Nicaragua had an acute mild COVID-19 infection that correlate with prolonged humoral response. This immune response to the RBD antigen, more likely IgG dependent, significantly increased between the acute to convalescent and decay in the late convalescent but still remained seropositive.

## INTRODUCTION

Coronavirus disease (COVID-19) is caused by the severe acute respiratory syndrome coronavirus 2 (SARS-CoV-2) discovered in Wuhan, China in mid-December 2019 [1]. The disease is characterized by fever, dry cough, shortness of breath, sore throat, myalgia, fatigue and headache, often accompanied by lymphopenia, prolonged prothrombin and elevated lactate dehydrogenase [2, 3]. Older people, and those with underlying medical conditions like cardiovascular disease, diabetes, chronic respiratory disease, and cancer are more likely to develop serious illness [4]. In contrast, young and healthy people infected with SARS-CoV-2 typically experience mild to moderate respiratory illness and recover without requiring special treatment [5, 6]. Despite the rapidly growing body of knowledge on SARS-CoV-2 infections, more information is needed to better understand disease susceptibility and immunity. For example, more information is needed on whether severe, moderate, mild or asymptomatic SARS-CoV-2 infection elicit persistent immune memory that is protective. The persistence of immunity has important implications for individual protection after infection and community immunity. Another priority for is understanding whether host genetic factors contribute to disease susceptibility or severity. In several studies, blood group O has been associated with lower risk of COVID-19, while non-O blood types are associated with increased risk [7].

The coronavirus (CoV) virion consists of a nucleocapsid core surrounded by an envelope containing three membrane proteins, spike (S), membrane (M) and envelope (E) that are common to all members of the genus [8]. SARS-CoV-2 is a β-CoV, a subgroup that includes SARS-CoV-1, MERS-CoV and the two common-cold human CoVs, OC43 and HKU-1 [9]. The Receptor Binding Domain (RBD) of the spike protein (S) is highly immunogenic and elicit antibodies that are strongly correlated with SARS-CoV-2 neutralization [10-12]. In longitudinal analyses SARS-CoV-2 IgA and IgM antibodies have been shown to decay rapidly, while IgG antibodies remained relatively stable up to 6 months or longer in serum and saliva [13-15]. The median time to seroconversion has been estimated to be between 9 and 12 days post-symptoms onset (PSO) for IgM, IgA and IgG antibodies against RBD [16, 17]. In contrast, the median times for IgM and IgA seroreversion appear to be around 49 and 71 days PSO, respectively [16].

Few SARS-CoV-2 longitudinal studies have been conducted in patients seeking care in the outpatients setting and in South American populations to estimate the burden of disease and to monitor the durability of immunity. Here we report on the burden of SARS-CoV-2 infections among patients with respiratory symptoms presenting to outpatient clinics during the first wave of the pandemic in Nicaragua. We also followed this cohort of patients for 6 months to assess the durability of SARS-CoV-2-specific antibodies among patients with an initial presentation of mild disease that resolved with no complications in the majority of the patients. Our results are applicable to estimating the burden of COVID-19 disease in Nicaragua and understanding the durability of antibodies among patients with mild to moderate disease.

## RESULTS

### Laboratory diagnosis of acute COVID-19 by RT-qPCR and paired serology

A total of 51 of the 157 outpatients had SARS-CoV-2 RNA detected in NP swabs collected during the acute phase; the Ct median was 24.6 (IQR, 20.3 – 32.0) (Table. 1). When paired acute and convalescent blood samples were tested for SARS-CoV-2 RBD specific antibody, we observed 47/157 (30%) of patients seroconverted confirming acute SARS-CoV-2 infection (Table. 1). In total 60 of 157 (38%) patients tested positive by PCR or paired serology (Fig. 1A). Of the 60 subjects diagnosed with acute SARS-CoV-2 infection, 38 (63%) were identified by both assays (Fig. 1B), 13 (22%) by RT-qPCR only (Fig 1C) and 9 (15%) by serology only (Fig.1D). Re-testing of the NP samples from subjects classified as “serology only” with a secondary, more sensitive, RT-qPCR assay showed amplification in 7 of 9, but Ct values were all >33 indicating low viral load. Of the 13 acute cases identified by PCR only, 5 subjects already had developed antibody in the acute specimen, with no evidence for seroconversion in those that provided a convalescent serum, 3 subjects did not develop detectable antibody in acute or convalescent specimens. The infection status of 5 subjects could not be established by serology as convalescent blood samples were not available (Fig. 1C). Of the 97 outpatients who tested negative for acute SARS-CoV-2 infection, 16 had RBD Specific-Ig antibody but not IgM in the acute specimen (collected <5 days PSO), which is indicative of a past infection not linked to the current illness (Fig. 1E). In summary, in the 157 patients evaluated, we observed 60 SARS-CoV-2 incident infections (38%) and 16 (10%) past infections.

**Table 1.** Summary of the RT-qPCR and Pan-Ig serology performed to confirm acute COVID-19 in outpatients with acute respiratory disease in Leon, Nicaragua between June and October 2021.

| Assay | Acute SARS-CoV-2 infection |  | Serology-positive on acute sample | Total |
| --- | --- | --- | --- | --- |
|  | Positive | Negative | Positive |  |
| RT-qPCR testing | 51 | 106 | - | 157 |
| Serology | 47<sup>a</sup> | 68 | 21 | 136<sup>b</sup> |
| **Final COVID-19 diagnosis** |  |  |  |  |
| Acute COVID-19 | 60 (38.0%) | 38 (63%) positive in both assays<br>13 (22%) positive by PCR only<br>5 convalescent samples unavailable for paired serology.<br>3 serology negative in acute and convalescent samples.<br>5 acute sample with positive serology<br>9 (15%) positive by serology only.<br>7 – tested positive in re-testing with modified primers |  |  |
| non-COVID-19 | 97 (62%) | 65 (67%) negative by both assays<br>5 of these seroconverted between visits 2 and 3<br>16 (16%) PCR negative and convalescent sample unavailable for paired serology.<br>16 (18 %) PCR negative and serology positive in acute sample, indicative of past infection. |  |  |
| Total | 157 (100%) |  |  |  |

<sup>a</sup> If the ratio of the convalescent/acute OD was $\geq 2$ and ratio $< 2$ but IgM-positive ( $OD \geq 0.300$ ) in serum collected after 5 days PSO.
<sup>b</sup> Acute and convalescent specimen were obtained from 132 of the 157 subjects enrolled in the study.

**Figure 1.**
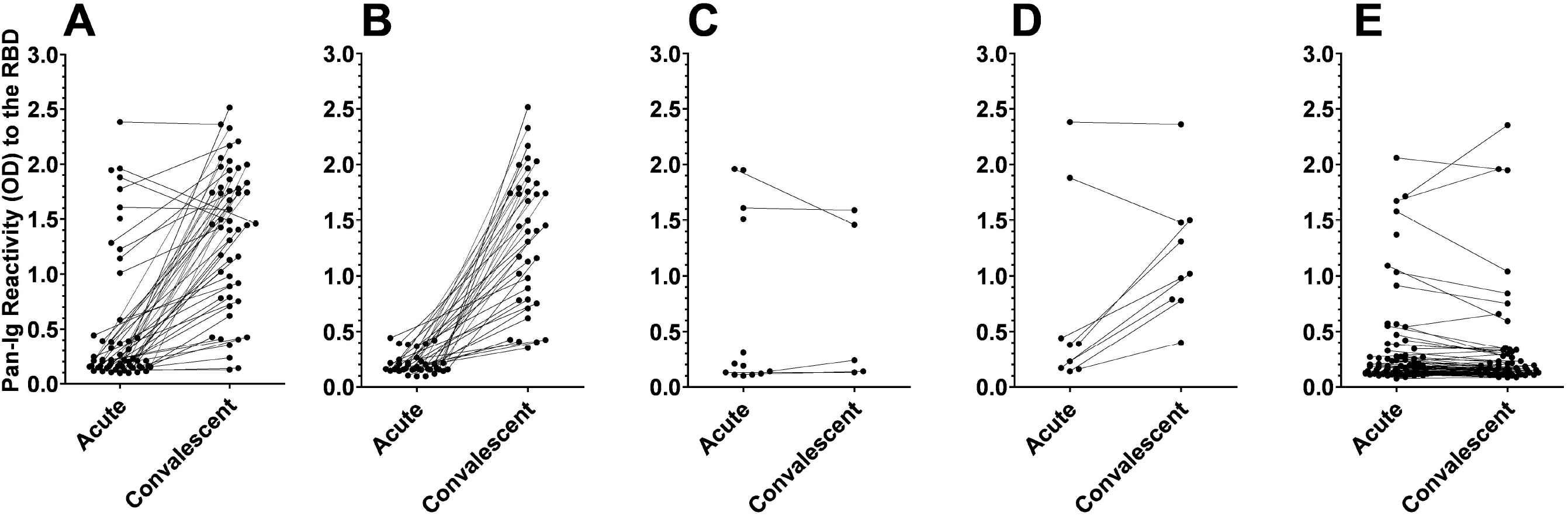
Patterns of Pan-Immunoglobulins response to the receptor binding domain of SARS-COV-2 in acute and convalescent serum from outpatients with acute COVID-19 diagnosed by RT-PCR and/or Serology between June and October 2020 in Nicaragua. A: All acute COVID-19 (n = 60), B: Acute-COVID-19 by RT-qPCR and Serology (n = 38), C: Acute COVID-19 by RT-qPCR only (n = 13), D: Acute-COVID by Serology only (n = 9) and E: All non-acute COVID-19 (n=97).

### Temporal patterns of reactivity to the SARS-RBD antigen

Subjects who tested seropositive in either acute or convalescent specimens were invited to provide a second late convalescent serum sample (>120 days PSO) to measure the durability of SARS-CoV-2 RBD binding antibodies. In total 53 subjects consented to participate, including 39 diagnosed with acute COVID-19 and 14 non-COVID-19 cases. The longitudinal analysis of RBD reactivity showed 3 different reactivity patterns. The first pattern observed in 29 subjects showed an increase in signal from acute to the first convalescent specimen and a decrease in signal from early and late convalescent specimen, likely due in part to the decline of IgA and IgM antibodies (Fig.2A and Fig 2E). The second pattern observed in 16 subjects showed a similar increase between the acute and the early convalescent but the signal remained high or even increased between the early and the late-convalescent specimens (Fig. 2B). The third pattern observed in 9 subjects showed either seroconversions (n = 5) or boost (n = 3) in antibody levels between the early and the late convalescent specimens indicative of incident infections and, possibly, re-exposure to antigen (Fig. 2C). Pan-Ig reactivity to the SARS-CoV-2 RBD antigen lasted for at least 6-month post-acute infection in the outpatients from this cohort (Fig. 2D) and IgG response was higher and more prolonged than IgM and IgA responses (Fig. 2E).

**Figure 2.**
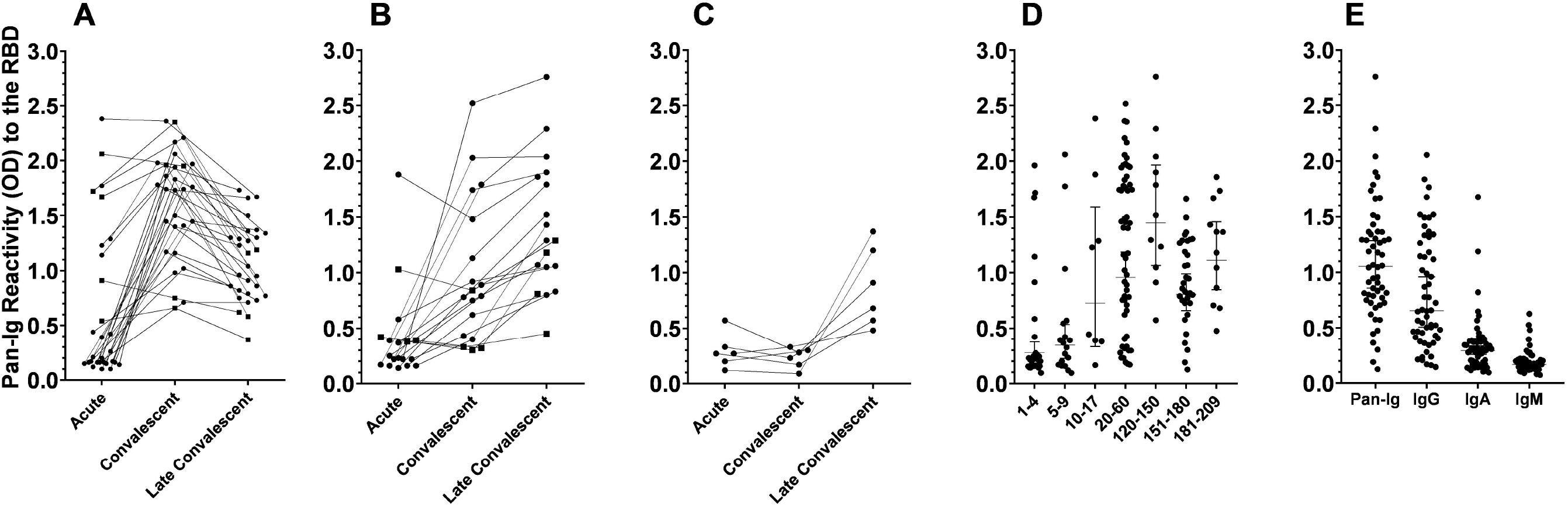
Reactivity to the SARS-CoV-2 RBD antigen over 6 months post symptoms onset in outpatients from Nicaragua. A: Subjects exposed to SARS-CoV-2 (n = 29), black dot represents acute COVID-19 and black square past infections. B: Subjects with non-decaying reactivity to the RBD antigen in late convalescent serum (n = 18). C: Subjects with either incident infection between convalescent and late serum (n = 6). D: correlation between Pan-Ig reactivity to the RBD and days PSO (n = 53). E: Correlation between Pan-Ig and isotypes reactivity to the RBD in late convalescent serum.

### Clinical and Epidemiological fingerprint of COVID-19 in outpatients from León, Nicaragua

Among several parameters examined in outpatient with acute respiratory disease, the following epidemiological characteristics were all associated (p < 0.05) with acute COVID-19 (n = 60) as compared to a non-COVID-19 illness (n = 97): age between 40 and 60 years (OR = 2.1), less frequent hand washing (≤5 times/day; OR = 2.8) and contact with a pet (OR = 2.8) (Table. 2). Among the clinical symptoms, fever (OR = 4.2), loss of taste (OR = 2.4), smell (OR = 3.0) and appetite (OR = 2.1) were associated with acute COVID-19 (Table. 3). The frequency of patients with ≥2 comorbidities was higher in acute-COVID-19 than in non-COVID-19 patients (28% vs 19%) and the most common was diabetes (18% vs 9%). After examination of several laboratory parameters, lower white cells count, presence of immature granulocytes, lower relative percent of eosinophils, higher INR, CRP-positivity and lower erythrocyte sedimentation rate were all associated with acute COVID-19 (Table. 4). Interestingly, while most hematologic parameters in convalescent blood normalized, participants with an acute COVID-19 diagnosis had persistently lower hemoglobin, red blood cell counts and hematocrit. Patients with acute COVID-19 were more likely to be treated with, ivermectin, azitromycin, aspirin and Vitamin C (Supplement). During the convalescent visit, 4 (2.5%) of 157 reported hospitalizations for their illness and 2 (1.3%) fatalities were confirmed; both of them had a diagnosis of acute COVID-19.

**Table 2.**
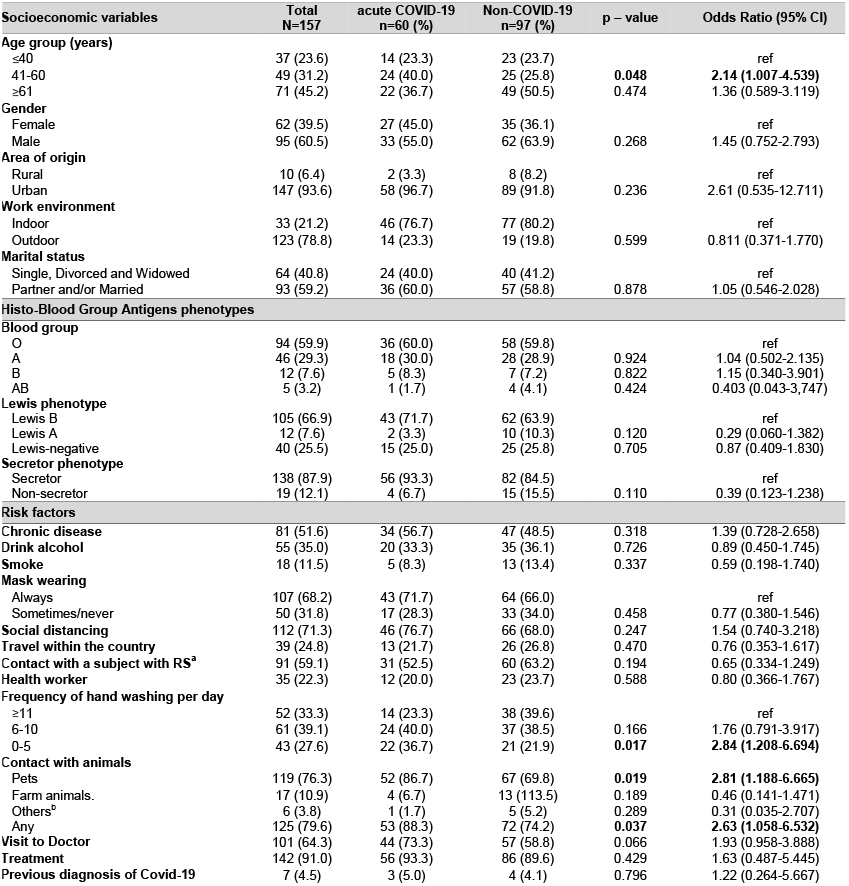
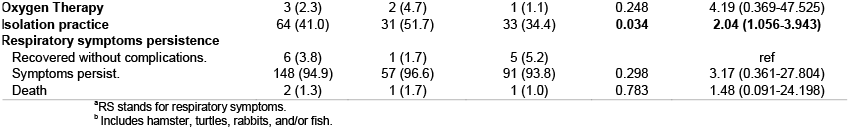
Correlation between epidemiological characteristics and SARS–CoV–2 infections in outpatients from Leon, Nicaragua. June-October 2020. N=157.

**Table 3.**
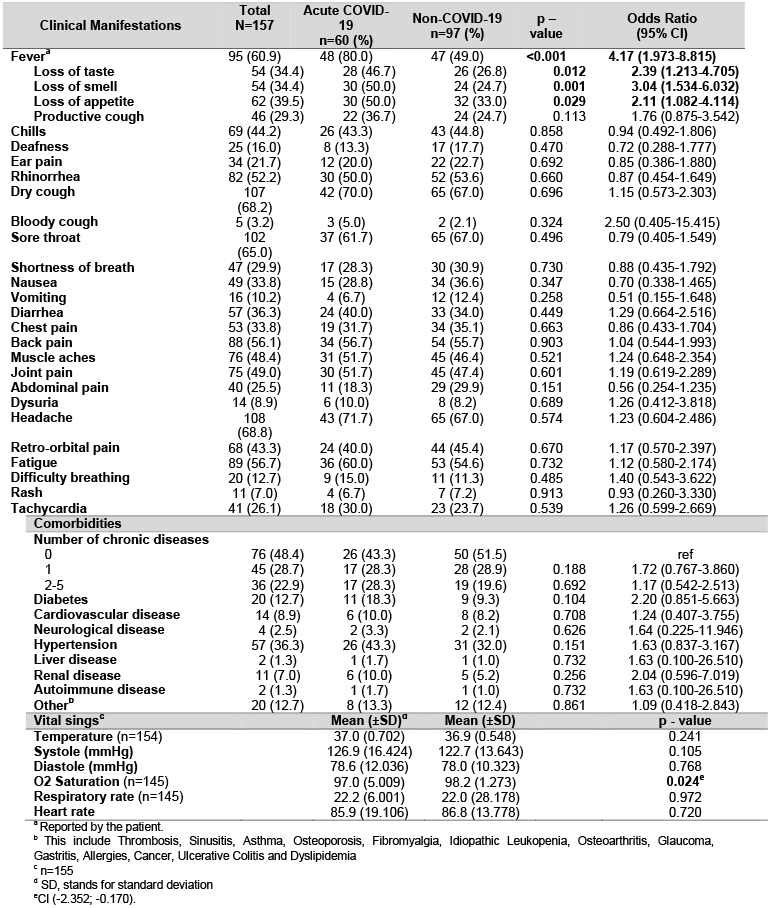
Clinical profile of SARS–CoV–2 infections in outpatients from Leon, Nicaragua. June-October 2020. N=157.

| Clinical Manifestations | Total<br>N=157 | Acute COVID-19<br>n=60 (%) | Non-COVID-19<br>n=97 (%) | p –<br>value | Odds Ratio<br>(95% CI) |
| --- | --- | --- | --- | --- | --- |
| Fever<sup>a</sup> | 95 (60.9) | 48 (80.0) | 47 (49.0) | <0.001 | 4.17 (1.973-8.815) |
| Loss of taste | 54 (34.4) | 28 (46.7) | 26 (26.8) | 0.012 | 2.39 (1.213-4.705) |
| Loss of smell | 54 (34.4) | 30 (50.0) | 24 (24.7) | 0.001 | 3.04 (1.534-6.032) |
| Loss of appetite | 62 (39.5) | 30 (50.0) | 32 (33.0) | 0.029 | 2.11 (1.082-4.114) |
| Productive cough | 46 (29.3) | 22 (36.7) | 24 (24.7) | 0.113 | 1.76 (0.875-3.542) |
| Chills | 69 (44.2) | 26 (43.3) | 43 (44.8) | 0.858 | 0.94 (0.492-1.806) |
| Deafness | 25 (16.0) | 8 (13.3) | 17 (17.7) | 0.470 | 0.72 (0.288-1.777) |
| Ear pain | 34 (21.7) | 12 (20.0) | 22 (22.7) | 0.692 | 0.85 (0.386-1.880) |
| Rhinorrhea | 82 (52.2) | 30 (50.0) | 52 (53.6) | 0.660 | 0.87 (0.454-1.649) |
| Dry cough | 107<br>(68.2) | 42 (70.0) | 65 (67.0) | 0.696 | 1.15 (0.573-2.303) |
| Bloody cough | 5 (3.2) | 3 (5.0) | 2 (2.1) | 0.324 | 2.50 (0.405-15.415) |
| Sore throat | 102<br>(65.0) | 37 (61.7) | 65 (67.0) | 0.496 | 0.79 (0.405-1.549) |
| Shortness of breath | 47 (29.9) | 17 (28.3) | 30 (30.9) | 0.730 | 0.88 (0.435-1.792) |
| Nausea | 49 (33.8) | 15 (28.8) | 34 (36.6) | 0.347 | 0.70 (0.338-1.465) |
| Vomiting | 16 (10.2) | 4 (6.7) | 12 (12.4) | 0.258 | 0.51 (0.155-1.648) |
| Diarrhea | 57 (36.3) | 24 (40.0) | 33 (34.0) | 0.449 | 1.29 (0.664-2.516) |
| Chest pain | 53 (33.8) | 19 (31.7) | 34 (35.1) | 0.663 | 0.86 (0.433-1.704) |
| Back pain | 88 (56.1) | 34 (56.7) | 54 (55.7) | 0.903 | 1.04 (0.544-1.993) |
| Muscle aches | 76 (48.4) | 31 (51.7) | 45 (46.4) | 0.521 | 1.24 (0.648-2.354) |
| Joint pain | 75 (49.0) | 30 (51.7) | 45 (47.4) | 0.601 | 1.19 (0.619-2.289) |
| Abdominal pain | 40 (25.5) | 11 (18.3) | 29 (29.9) | 0.151 | 0.56 (0.254-1.235) |
| Dysuria | 14 (8.9) | 6 (10.0) | 8 (8.2) | 0.689 | 1.26 (0.412-3.818) |
| Headache | 108<br>(68.8) | 43 (71.7) | 65 (67.0) | 0.574 | 1.23 (0.604-2.486) |
| Retro-orbital pain | 68 (43.3) | 24 (40.0) | 44 (45.4) | 0.670 | 1.17 (0.570-2.397) |
| Fatigue | 89 (56.7) | 36 (60.0) | 53 (54.6) | 0.732 | 1.12 (0.580-2.174) |
| Difficulty breathing | 20 (12.7) | 9 (15.0) | 11 (11.3) | 0.485 | 1.40 (0.543-3.622) |
| Rash | 11 (7.0) | 4 (6.7) | 7 (7.2) | 0.913 | 0.93 (0.260-3.330) |
| Tachycardia | 41 (26.1) | 18 (30.0) | 23 (23.7) | 0.539 | 1.26 (0.599-2.669) |
| **Comorbidities** |  |  |  |  |  |
| **Number of chronic diseases** |  |  |  |  |  |
| 0 | 76 (48.4) | 26 (43.3) | 50 (51.5) |  | ref |
| 1 | 45 (28.7) | 17 (28.3) | 28 (28.9) | 0.188 | 1.72 (0.767-3.860) |
| 2-5 | 36 (22.9) | 17 (28.3) | 19 (19.6) | 0.692 | 1.17 (0.542-2.513) |
| Diabetes | 20 (12.7) | 11 (18.3) | 9 (9.3) | 0.104 | 2.20 (0.851-5.663) |
| Cardiovascular disease | 14 (8.9) | 6 (10.0) | 8 (8.2) | 0.708 | 1.24 (0.407-3.755) |
| Neurological disease | 4 (2.5) | 2 (3.3) | 2 (2.1) | 0.626 | 1.64 (0.225-11.946) |
| Hypertension | 57 (36.3) | 26 (43.3) | 31 (32.0) | 0.151 | 1.63 (0.837-3.167) |
| Liver disease | 2 (1.3) | 1 (1.7) | 1 (1.0) | 0.732 | 1.63 (0.100-26.510) |
| Renal disease | 11 (7.0) | 6 (10.0) | 5 (5.2) | 0.256 | 2.04 (0.596-7.019) |
| Autoimmune disease | 2 (1.3) | 1 (1.7) | 1 (1.0) | 0.732 | 1.63 (0.100-26.510) |
| Other<sup>b</sup> | 20 (12.7) | 8 (13.3) | 12 (12.4) | 0.861 | 1.09 (0.418-2.843) |
| **Vital sings<sup>c</sup>** |  | **Mean (±SD)<sup>d</sup>** | **Mean (±SD)** | **p - value** |  |
| Temperature (n=154) |  | 37.0 (0.702) | 36.9 (0.548) | 0.241 |  |
| Systole (mmHg) |  | 126.9 (16.424) | 122.7 (13.643) | 0.105 |  |
| Diastole (mmHg) |  | 78.6 (12.036) | 78.0 (10.323) | 0.768 |  |
| O2 Saturation (n=145) |  | 97.0 (5.009) | 98.2 (1.273) | 0.024<sup>e</sup> |  |
| Respiratory rate (n=145) |  | 22.2 (6.001) | 22.0 (28.178) | 0.972 |  |
| Heart rate |  | 85.9 (19.106) | 86.8 (13.778) | 0.720 |  |

<sup>a</sup> Reported by the patient.
<sup>b</sup> This include Thrombosis, Sinusitis, Asthma, Osteoporosis, Fibromyalgia, Idiopathic Leukopenia, Osteoarthritis, Glaucoma, Gastritis, Allergies, Cancer, Ulcerative Colitis and Dyslipidemia
<sup>c</sup> n=155
<sup>d</sup> SD, stands for standard deviation
<sup>e</sup> CI (-2.352; -0.170).

**Table 4.** Correlation between blood parameters and SARS-CoV-2 infection in outpatients from Leon, Nicaragua, June to October 2021. (N=157)

| Laboratory indicators<br>(acute) | acute COVID-19<br>n=60<br>Mean (±SD)<sup>a</sup> | non-COVID-19<br>n=97<br>Mean (±SD) | p - value |
| --- | --- | --- | --- |
| White blood cell count (cells/μl) | 6.4x10<sup>3</sup> (2.2 x10<sup>3</sup>) | 8.1x10<sup>3</sup> (2.4 x10<sup>3</sup>) | **0.000\*** |
| Neutrophil (%) | 57.9 (11.526) | 58.7 (12.369) | 0.656 |
| Immature Granulocyte (%) | 0.10 (0.354) | 0.00 (0) | **0.006\*\*** |
| Eosinophil (%) | 1.05 (1.661) | 1.97 (2.033) | **0.004\*\*\*** |
| Basophil (%) | 0.43 (0.810) | 0.56 (0.957) | 0.389 |
| Lymphocyte (%) | 35.5 (11.158) | 34.6 (11.759) | 0.618 |
| Monocyte (%) | 5.0 (3.998) | 4.1 (2.734) | 0.084 |
| Platelets/μl | 2.1x10<sup>5</sup> (5.5 x10<sup>4</sup>) | 2.2x10<sup>5</sup> (6.0 x10<sup>4</sup>) | 0.153 |
| Red blood cells/μl | 4.5x10<sup>6</sup> (3.9 x10<sup>6</sup>) | 4.4x10<sup>6</sup> (4.4 x10<sup>6</sup>) | 0.673 |
| Hematocrit, % | 40.3 (3.565) | 39.8 (3.927) | 0.631 |
| Hemoglobin (g/dl) | 13.4 (1.191) | 13.3 (1.304) | 0.770 |
| Prothrombin time (seconds)<sup>b</sup> | 14.5 (2.770) | 14.4 (2.489) | 0.776 |
| Partial thromboplastin time (seconds)<sup>b</sup> | 42.3 (17.034) | 45.6 (21.685) | 0.490 |
| International normalized ratio (INR)<sup>b</sup> | 1.29 (0.316) | 1.13 (0.163) | **0.009\*\*\*\*** |
| ESR in men<sup>c</sup> | 11.4 (7.11) | 15.7 (8.89) | **0.040\*\*\*\*\*** |
| ESR in women<sup>d</sup> | 17.0 (9.67) | 15.5 (10.71) | 0.500 |
| C reactive protein (CRP) positive<sup>e</sup> | 42 (70.0) | 48 (50.0) | **0.015\*\*\*\*\*** |
| CRP concentration (mg/L) |  |  |  |
| 6-23 | 30 (71.4) | 25 (52.1) | ref |
| 24-196 | 12 (28.6) | 23 (47.9) | 0.063 |
| Abnormal parameters in urine<sup>ef</sup> | 9 (15.3) | 14 (15.4) | 0.983 |

| Laboratory indicators<br>(convalescent) | Acute-COVID-19<br>n=55<br>Mean (±SD) | non-COVID-19<br>n=81<br>Mean (±SD) | p - value |
| --- | --- | --- | --- |
| White blood cell count | 9.1x10<sup>3</sup> (3.3 x10<sup>3</sup>) | 8.6x10<sup>3</sup> (2.5 x10<sup>3</sup>) | 0.417 |
| Neutrophil | 57.0 (9.229) | 57.4 (8.897) | 0.819 |
| Eosinophil | 1.96 (2.160) | 2.23 (2.373) | 0.492 |
| Basophil | 0.45 (0.789) | 0.54 (0.807) | 0.525 |
| Lymphocyte | 37.0 (8.754) | 37.5 (9.886) | 0.760 |
| Monocyte | 3.45 (3.276) | 2.70 (2.619) | 0.159 |
| Platelet count | 2.5x10<sup>5</sup> (7.6 x10<sup>4</sup>) | 2.6x10<sup>5</sup> (6.5 x10<sup>4</sup>) | 0.233 |
| Hemoglobin, g/ml | 12.7 (1.208) | 13.2 (1.252) | **0.013##** |
| Red blood cell count | 4.2x10<sup>6</sup> (4.1 x10<sup>5</sup>) | 4.4x10<sup>6</sup> (4.3 x10<sup>5</sup>) | **0.006###** |
| Hematocrit, % | 38.2 (3.621) | 40.0 (4.043) | **0.007####** |
| ESR in men | 17.8 (8.01) | 13.8 (7.12) | 0.068 |
| ESR in women | 20.5 (10.39) | 18.7 (11.33) | 0.440 |

<sup>a</sup> SD, stands for standard deviation
<sup>b</sup> Examined in 25 acute COVID-19 and in 46 non-COVID-19.
<sup>c</sup> ESR stands for Erythrocyte sedimentation rate and was examined in 27 acute COVID-19 and 34 non-COVID-19
<sup>d</sup> Examined in 32 acute COVID-19 and 62 non-COVID-19
<sup>e</sup> For this parameter the number represent frequency and percentages in parenthesis, instead of mean and SD.
<sup>f</sup> The presence of nitriles, while blood cells and bacteria were considered abnormal.
\*CI (-2459.947; -941.015); \*\*CI (0.029; 0.171); \*\*\*CI(-1.536; -0.303); \*\*\*\*CI(0.038; 0.265); \*\*\*\*\*OR 2.33, CI(1.180-4.614); \*\*\*\*\* CI(-8.407;-0.205); ## CI (-0.9673; -0.1139); ###CI(-350098.4; -60653.5); ####CI(-3.1586; -0.4777);

There was an association between the frequency of outpatient with diagnosis of acute COVID-19 from the current study and confirmed cases from Nicaragua, mainly derived from hospitalized patients, reported in the Johns Hopkins University website [18] (Fig. 3).

**Figure. 3.**
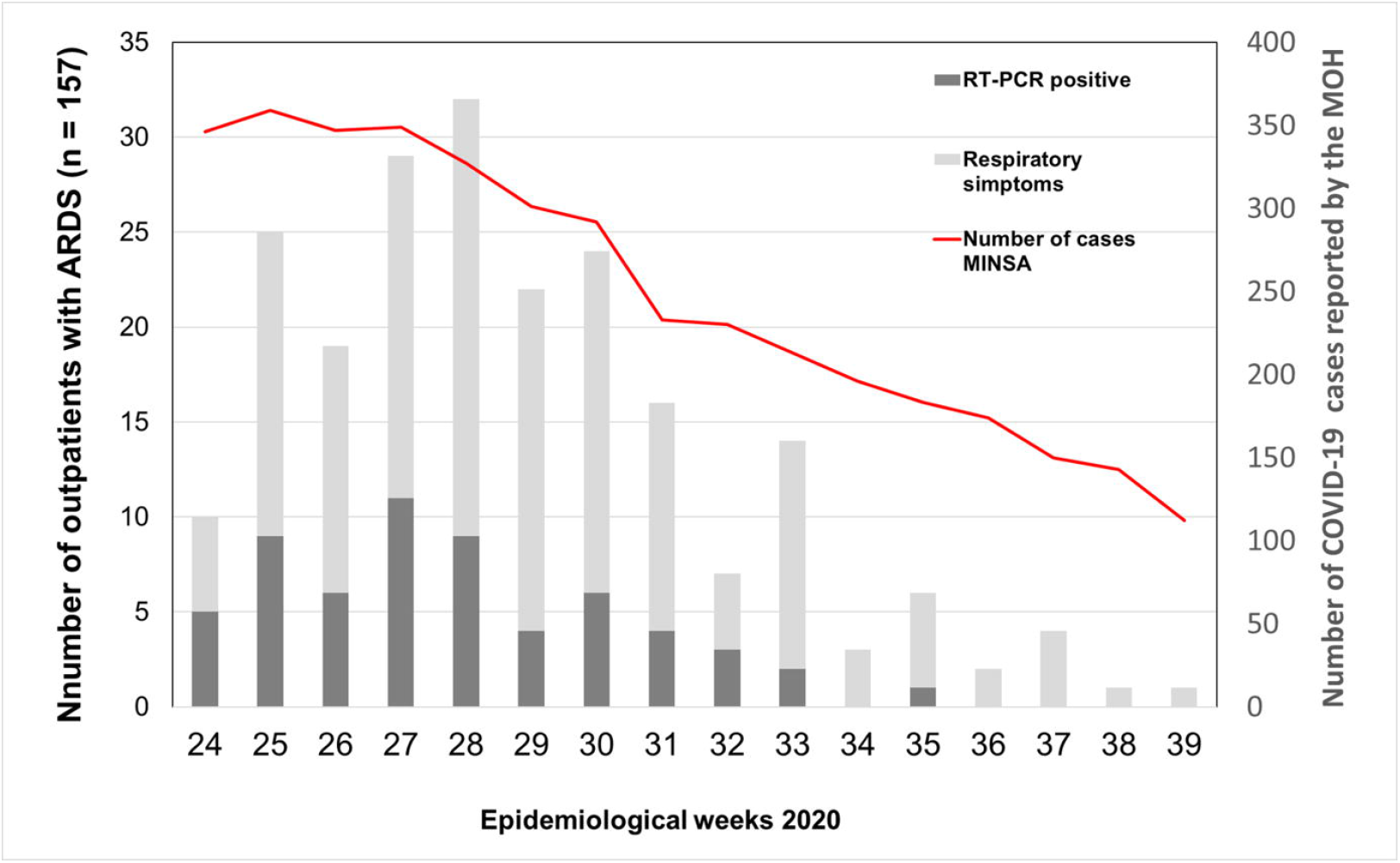
Correlation between the frequency of outpatients with acute COVID-19 from the current study and the reported in the Johns Hopkins University website to track COVID-19 in real time which is according to the Ministry of Health from Nicaragua.

## DISCUSION

The current study contributes to our understanding of the humoral response to SARS-CoV-2 over 6 months in outpatients in Nicaragua a country with limited epidemiological and immunological data during the COVID-19 pandemic. This study also extends previous knowledge on clinical, epidemiological, laboratory diagnosis and treatment associated with acute SARS-CoV-2 infection in outpatients.

Molecular and serological testing confirmed that more than one third of the outpatients seeking care for acute respiratory disease during the first epidemic wave of SARS-CoV-2 in Nicaragua had an acute COVID-19 infection (Table 1). Other infectious etiologies with a similar clinical and epidemiological profile that may have been co-circulating in this setting at the same time frame (June – October 2020), such as influenza or pneumococcal, remain to be investigated. Acute respiratory infections typically peak in incidence between July and October in Nicaragua [19].

Initial serologic analysis to explore antibody response to the RBD antigen showed 3 categories, the first includes subjects with naïve acute serum but highly reactive convalescent serum to the RBD antigen (Fig. 1B), the second were subjects with acute and convalescent serum highly or mildly reactive to the RBD antigen, indicative of prior infection if the serum was collected < 5 days PSO (Fig. 1E) and the third includes subjects that do not experience seroconversion to the RBD antigen despite RNA detection in NP by RT-qPCR (Fig. 1C). The reactivity to the RBD antigen observed in most of the acute outpatients with COVID-19 indicate activation of the humoral response and is suggestive of the development of neutralizing antibodies with the capacity to block SARS-COV-2 spike protein interaction with its cognate receptor ACE-2 as previously reported [17, 20]. The presence of viral RNA without serological response suggests false-positive PCR or atypical antibody responses, which have been reported in some patients [17, 21]. Genomic studies have shown a continuous emergence of novel variants in several populations, although there is little information on the presence of these or other variants in Central American populations [22, 23]. It was also interesting to observe RT-qPCR negative individuals with strong reactivity to the RBD antigen in convalescent serum suggestive of false-negative RT-qPCR test (Fig. 1D). Re-examination with an alternative RT-qPCR that used more sensitive primers and probes to the E region [24] than the Charité system [25] showed RNA amplification in 7 of those 9 subjects, confirming that these were indeed false negatives. Our results demonstrate the value of using both RT-qPCR and paired serology for laboratory confirmation of acute COVID-19 cases.

We also analyzed a subset of acute COVID-19 cases to understand the long-term dynamics of antibody responses. We observed three patterns of changes in antibody levels over a 6 month follow up period. In the first pattern, reactivity to the RBD antigen significantly increased between the acute to convalescent, thereafter decreased in the late convalescence, but remained seropositive up to 6 months (Figs. 2A). In the second pattern, the reactivity did not decay and remained high between convalescent and late convalescent sera, indicative of strong immune response over 6 months (Fig. 2B). The third pattern was characterized by incident infections and boost in the reactivity to RBD between convalescent and late convalescent serum. The boost in Ab levels between convalescence and late convalescence may indicate a secondary subclinical infection, possibly by SARS-CoV-2 variants.

The clinical symptoms and laboratory profiles of this Nicaraguan outpatient cohort follows the trend observed in hospitalized patients from several other studies [27, 28]. This profile includes loss of taste, smell and appetite, age 40 to 60 years, contact with pets, reduced hand washing, lower leukocytes counts, lower eosinophils, increased INR and increased erythrocyte sedimentation rate. As compared with non-COVID-19 patients, C-reactive protein (CRP) test was more likely to be positive in COVID-19 outpatients (42% vs 48%, *p* < 0.05), suggesting that CRP positivity is common in patients mild COVID-19 disease as observed in severe cases with excessive inflammation [29]. In previous studies, the degree of lymphocytes count gives a hint for disease prognosis and is found to be positively correlated with disease severity [27]. For instance, preliminary studies from Wuhan reported lymphopenia in 80% of the critical patients and 25% in patients with mild COVID-19 infection [30, 31]. Given the significantly lower leucocytes counts observed in COVID-19 patients in this study, is likely that they also experienced lower lymphocytes counts, but with similar differential white blood cell proportions (35.5% vs 34.6%) as compare with non-COVID-19 patient, which is in line with mild disease. It has previously been suggested that lower eosinophils counts is associated with acute respiratory deterioration during SARS-CoV-2 [32], the current study further show that lower eosinophils counts is also associated with non-severe disease and might be considered a marker of infection in general. A similar multidrug therapy (ivermectin, azithromycin and aspirin) to the used in the current study have shown to improve recovery and prevent risk of hospitalization and death among ambulatory COVID-19 cases in Mexico [33], but further effectiveness studies will be needed to reach any conclusion.

The correlation between the COVID-19 status and the distribution of ABO, Lewis and Secretors phenotypes was explored, but the lower frequency of the non-O blood, LeA and Se-phenotypes in this population required a higher sample size to establish a reliable conclusion.

In conclusion, this study report that more than one third of the outpatients seeking care for acute respiratory disease during the first epidemic wave of SARS-CoV-2 in Nicaragua had an acute mild COVID-19 infection that correlate with prolonged humoral response. This immune response to the RBD antigen, more likely IgG dependent, significantly increased between the acute to convalescent and decay in the late convalescent but still remained seropositive.

## MATERIAL AND METHODS

### Study subjects

A total of 157 patients, with a median of 43 years of age (IQR, 32 - 59), seeking care for respiratory disease suggestive of COVID-19 in private healthcare clinics were prospectively enrolled between June to October 2020 in Leon, Nicaragua. Following the clinician’s consultation each patient was asked by phone to participate in the study. Interested patients were visited in their household by the study team within 17 days PSO to obtain informed consent and collect clinical and demographic data in a questionnaire. Nasopharyngeal swabs and blood samples were collected for RT-qPCR and Pan-immunoglobulin (IgM, IgG and IgA) screening, respectively. Patients were also asked to provide a second blood sample during convalescent phase, 20 to 60 days PSO, and a third blood sample (late-convalescent) between 120 – 180 days PSO. This study was approved by the ethical committee for biomedical research from UNAN-León, on May 2020 and amended in September 2020 (FWA00004523/IRB00003342). All patients signed an inform consent and all methods applied in accordance with guidelines and regulations, such as, good clinical and laboratory practices.

### Samples collection

Nasopharyngeal samples (NP) were collected following the recommendations by the World Health Organization (WHO) for the rational use of personal protective equipment (PPE) in health care and home care settings [34].

### Assays for examination hematological parameters and markers of inflammation

The relative percentages of neutrophil, lymphocyte, eosinophil, monocyte and basophil were manually examined by using Wright staining. Qualitative and semi-quantitative C reactive protein (CRP), partial thromboplastin time (PTT) and prothrombin time (PT) were determined with the HumaTex CRP, Hemostat aPTT-EL and Hemostat Thromboplastin-SI latex agglutination kits, respectively (Human, Wiesbaden Germany). ABO blood groups were determined by haemagglutination (Cypress Diagnostics, Hulshout, Belgium). Lewis A (LeA) Lewis B (LeB) and H (secretor) antigens were detect using an in-house saliva-based ELISA [35].

### RT-qPCR for SARS-CoV-2 screening

Viral RNA extraction was performed from 140μl of the solution containing the nasopharyngeal (NP) swab by using the QIAamp Viral RNA Mini Kit (Qiagen, Hilden, Germany), according to manufacturer’s instructions. Purified viral RNA was analyzed by RT-qPCR for SARS-CoV-2 screening by using the Sarbeco-E primers and probes described in the Charité assay [25]. The primers and probes for the RdRp gene described in the Charité assay were not considered based in a previous report showing low sensitivity [36]. RT-qPCR was performed with the AgPath-ID OneStep RT-PCR Kit (Thermo Fisher Scientific, Waltham, MA) using a Light Cycler®96 (Roche, Mannheim, Germany). A sample was considered positive if the cycle threshold (Ct) value was ≤ 36. NP samples from patients diagnosed by serology only were re-examined with primers and probes described by Smyrlaki and coworkers, Ct ≤ 36 [24].

### In-house RBD ELISA

The protein expression and purification of the recombinant SARS-CoV-2 RBD antigen used were previously described by Premkumar and coworkers [17]. The sensitivity and specificity of this assay to detect SARS-CoV-2 antibodies have been found to be 98% and 100% at 9 days PSO, respectively [17]. In brief, anti-RBD specific immunoglobulins (Ig) and IgM antibodies (ab) were determined by ELISA using heat inactivated serum at 56°C for 30 minutes. The 96-well high-binding microtiter plate (Greiner bio one cat # 655061) was coated with 50 µl of the RBD antigen at 4µg/ml in Tris Buffered Saline (TBS) pH 7.4 for 1 hr at 37°C. Plates were blocked with 100 µl of blocking solution (3% non-fat milk (Pinito) in TBS containing 0.05% Tween 20) for 1 hr at 37°C and 50 µl of serum at 1:40 in blocking buffer was added and incubated for 1 hr at 37°C. After washing (TBS-tween-tween 0.2% v/v), 50 µl of goat anti-human alkaline phosphatase conjugated antibody mixture at 1:2500 dilution was added for 1 hr at 37°C 1. The mixture contains anti-IgG (Sigma Cat # A9544), anti-IgA (Ab cam Cat # AB97212), and anti-IgM (Sigma Cat # A3437) antibodies conjugated with alkaline phosphatase. After washing, 50 µl P-Nitrophenyl phosphate substrate (SIGMA FAST, Cat No N2770) was added to the wells and the optical density (OD) was measured after 10 min at 405nm by using a plate reader (Biotek Epoh). Acute serum with OD readings of ≥0.300 in the Pan-immunoglobulin RBD assay (Pan-Ig-RBD) were defined as seropositive. OD readings of ≥ 0.300 in acute samples collected ≤5days PSO were indicative of prior infection if Pan-Ig was positive but IgM-negative. In seropositive patients, a recent infection or seroconversion was defined as the ratio between the OD-convalescent/OD-acute ≥2. All subjects seropositive in acute and convalescent serum were asked to provide a third sample to investigate prolonged humoral response. All acute, convalescent and late convalescent serum samples were analyzed for IgG, IgA and IgM separately using isotype specific goat anti-human antibody by following the same procedure used for Pan-Ig.

### Statistics

Patients RT-qPCR-positive and/or seroconverted were defined as acute COVID-19 and RT-qPCR-negative with non-seroconversion was defined as non-COVID-19. Arithmetic mean with standard deviation (±SD) was used to compare continuous variables. Frequency and percentages were used to compare categorical variables. The differences between two groups of continuous variables was tested by using independent *t-test*. A regression univariate model was applied to explore risk factors associated with acute COVID-19, only categorical variables were included in the model. Odds Ratio (OR) was calculated to determine the degree of association at 95% confident interval (95%, CI), the association was significant if α was <0.05, all statistical tests were two-sided. All statistical analysis was performed in Statistical Package for the Social Sciences (SPSS version 21).

## Data Availability

The authors confirm that the data supporting the findings of this study are available within the article [and/or] its supplementary materials.

## ACKNOWLEDGEMENTS

Authors would like to thanks all outpatients in the current study for attending the fieldwork staff and for providing the clinical samples. We also appreciate the support from Dr. Fabian Padilla for the identification of COVID-19 suspected patients. FG, OZ, and YR are supported by an award from the NIH-Fogarty International Center [D43TW010923]. SBD and LP are supported by awards from the National Institute of Allergy and Infectious Diseases [K24AI141744 and 1U01AI151788-01, respectively]. LP also receive support from the National Cancer Institute [NCI U54 CA260543-01] FB, JN and LS received support from the Swedish Research Council.

## AUTHORS CONTRIBUTION

All authors have reviewed and approved this manuscript. Personal contribution are as follows: FG, OZ, EC and YR carried out the experiments and contribute with data analysis, CTR contribute with statistical analysis, AM carried out the clinical evaluation of the patients and review the clinical data, HV and NM collected the clinical and epidemiological information, collected the biological material and perform experiments, LS and JN contribute in the study design and data interpretation, AMS and LP synthetize de reagents, contribute with the study design and perform data and manuscript curation, SBD contribute with study design, statistical analysis revision and participate in the manuscript preparation. FB contribute with study design, perform data analysis, coordinate laboratory and field activities and wrote the manuscript.

