## Supplemental 1 for "Antibody response to SARS-CoV-2 infection over six months among Nicaraguan outpatients"

| Treatment |  | Azithromycin | Ivermectin | Aspirin | Vitamins | Colchicine | Ibuprofen | Acetaminophen | Anti-inflammatories | Cough and cold medicines | Home remedies | Other antibiotics | Others  Treatment |
| --- | --- | --- | --- | --- | --- | --- | --- | --- | --- | --- | --- | --- | --- |
| Azithromycin (%) | Acute –Covid-19^€^ | 56.7 |  |  |  |  |  |  |  |  |  |  |  |
|  | Non-Covid19^¥^ | 42.3 |  |  |  |  |  |  |  |  |  |  |  |
| Ivermectin (%) | Acute –Covid-19 | 40.0 | 48.3 |  |  |  |  |  |  |  |  |  |  |
|  | Non-Covid19 | 25.8 | 32.0 |  |  |  |  |  |  |  |  |  |  |
| Aspirin (%) | Acute –Covid-19 | 23.3 | 16.7 | 25.0 |  |  |  |  |  |  |  |  |  |
|  | Non-Covid19 | 10.3 | 8.2 | 14.4 |  |  |  |  |  |  |  |  |  |
| Vitamins^β^ (%) | Acute –Covid-19 | 20.0 | 15.0 | 6.7 | 26.7 |  |  |  |  |  |  |  |  |
|  | Non-Covid19 | 7.2 | 6.2 | 1.0 | 13.4 |  |  |  |  |  |  |  |  |
| Colchicine (%) | Acute –Covid-19 | 18.3 | 20.0 | 10.0 | 6.7 | 23.3 |  |  |  |  |  |  |  |
|  | Non-Covid19 | 6.2 | 8.2 | 2.1 | 2.1 | 8.2 |  |  |  |  |  |  |  |
| Ibuprofen (%) | Acute –Covid-19 | 15.0 | 13.3 | 8.3 | 10.0 | 5.0 | 23.3 |  |  |  |  |  |  |
|  | Non-Covid19 | 10.3 | 7.2 | 2.1 | 2.1 | 2.1 | 18.6 |  |  |  |  |  |  |
| Acetaminophen (%) | Acute –Covid-19 | 13.3 | 13.3 | 8.3 | 5.0 | 1.7 | 5.0 | 26.7 |  |  |  |  |  |
|  | Non-Covid19 | 10.3 | 5.2 | 6.2 | 0.0 | 2.1 | 2.1 | 19.6 |  |  |  |  |  |
| Anti-inflammatories^α^ (%) | Acute –Covid-19 | 11.7 | 11.7 | 5.0 | 13.3 | 5.0 | 10.0 | 1.7 | 23.3 |  |  |  |  |
|  | Non-Covid19 | 7.2 | 5.2 | 1.0 | 3.1 | 1.0 | 2.1 | 2.1 | 12.4 |  |  |  |  |
| Cough and cold medicines^µ^ (%) | Acute –Covid-19 | 11.7 | 11.7 | 3.3 | 3.3 | 8.3 | 5.0 | 5.0 | 3.3 | 18.3 |  |  |  |
|  | Non-Covid19 | 5.2 | 4.1 | 2.1 | 0.0 | 1.0 | 3.1 | 4.1 | 0.0 | 8.2 |  |  |  |
| Home remedies (%) | Acute –Covid-19 | 6.7 | 5.0 | 1.7 | 3.3 | 5.0 | 1.7 | 3.3 | 5.0 | 0.0 | 11.7 |  |  |
|  | Non-Covid19 | 10.3 | 7.2 | 2.1 | 2.1 | 2.1 | 3.1 | 5.2 | 2.1 | 1.0 | 17.5 |  |  |
| Other antibiotics^π^ (%) | Acute –Covid-19 | 3.3 | 5.0 | 1.7 | 3.3 | 1.7 | 1.7 | 5.0 | 3.3 | 1.7 | 0.0 | 11.7 |  |
|  | Non-Covid19 | 1.0 | 4.1 | 0.0 | 1.0 | 1.0 | 0.0 | 2.1 | 1.0 | 1.0 | 0.0 | 10.3 |  |
| Others Treatment^Ω^ (%) | Acute –Covid-19 | 23.3 | 23.3 | 6.7 | 15.0 | 13.3 | 10.0 | 13.3 | 11.7 | 6.7 | 6.7 | 5.0 | 45.0 |
|  | Non-Covid19 | 24.7 | 17.5 | 8.2 | 9.3 | 6.2 | 9.3 | 7.2 | 7.2 | 2.1 | 6.2 | 6.2 | 47.4 |

Table 5. Correlation between the pharmacological treatment and COVID-19 status in outpatients attending private clinics of León, Nicaragua. June – October 2021.

^€^Acute –Covid-19 N=60

^¥^ Non-Covid19 N=97

β Included mainly vitamin C followed by D in minor proportion

α Included Dexamethasone, Hydrocortisone, Apronax, Dolofin, Diclofenac, Novalgin, Deflazacort

µ Included Asprax, Umquan, Prednisone, sudagrip, I.R.S, Ambroxol, RIMOSS Nasal spray, Menaxol, Actimycin, Abrilar, Dologrip

π Included Uvamin, Ciprofloxacin, Cistericin, Levofloxacin, Moxiflox, Cefixime, Ceftriaxone, Trimethopin Sulfamethoxazole

Ω Included Hepacequel, Calcium, Decathylene, Cardioaspirin, Omeprazole, Relax Plus, Ultra Doceplex, Thiamine Forte, Probiotics, Magnesium, Antihistamines, Beclomethasone, Viusid, Enoxaparin, Lansoprazole, Nitazoxanide, Metronidazole, Pulmicort, Sodium Sulfate, Hemihydrate, Iron, Cetriler, Phenylephrine, Hydroxyzine, Isosorbide, Cetirizine, Enalapril, Salbutamol with Beclomethasone, Albuterol, Validal, Magnum, Lepirel, Acyclovir, Loratadine
